# Life’s Crucial 12: Updating and Enhancing the Life’s Essential 8 of Cardiovascular Health: A proposal from NHANES

**DOI:** 10.1101/2024.05.08.24307090

**Authors:** Ruoyu Gou, Yufan Gou, Danni Dou, Guanghua Li

**Author notes:** Correspondence: Guanghua Li. These authors have the same contribution (Ruoyu Gou, Yufan Gou, Danni Dou).

## Abstract

**Background:** Life’s Essential 8 (LE8) is a cardiovascular health (CVH) model but does not take into account mental health, an important cardiovascular risk factor, so we constructed Life’s Crucial 12 (LC12), a comprehensive cardiovascular care model that takes CVH into account, based on LE8, and hypothesized that it would be a more reliable index of CVH, despite the additional information needed to calculate LC12.

**Objective:** To construct an integrated cardiovascular care model LC12 based on LE8 that can take Psychological Health into account, and to report the association between LC12 and stroke.

**Design:** Population-based, cross-sectional study.

**Setting:** Various locations in the United States.

**Participants:** This study was a cross-sectional study based on data from the 2005-2008 National Health and Nutrition Examination Survey (NHANES), which included 4,478 U.S. adults (≥ 20 years old).

**Method:** The composite cardiovascular care model LC12 with scores (range 0-100) defining low (0-49), medium (50-79) and high (80-100) CVH. Determination of stroke status was obtained by questionnaire. Associations were assessed using multivariate logistic regression models and restricted cubic spline models.

**Result:** Among 4,478 participants, there were 2252 female and 2226 male participants (53.136% and 46.864%, respectively), and 250 participants (5.583%) were diagnosed with stroke. The mean values of LC12, Psychological Health, Health behaviors, and Health factors scores for participants with stroke were 68.953, 52.775, and 55.451, respectively, which were lower than those of Non-Stroke participants. After fully adjusting for confounders, the ORs for the LC 12, Psychological Health, Health Behaviors, and Health Factors moderate and high groups were 0.431 (0.226,0.822), 0.212 (0.060,0.755), 0.536 (0.297, 0.967), 0.357 (0.178,0.713), 0.759 (0.552, 1.043), 0.334 (0.179, 0.623), 0.565 (0.406, 0.786), 0.533 (0.286, 0.994), which were significantly associated with the risk of stroke (*P-trend* < 0.05) and there was a linear trend between subgroups with different scores (*P-value* < 0.001). However, no nonlinear dose relationship was observed (*P-Nonlinearity* > 0.05).

**Limitation:** Because estimates are based on single measures, fluctuations over time could not be determined.

**Conclusion:** These findings suggest that Psychological Health is important in CVH. CVH status assessed by LC12 (Psychological Health, Health behaviors, Health factors) was significantly associated with the risk of developing stroke. When LC12 scores are maintained at high levels, it is beneficial to decrease the risk of stroke.

**Abstract Pictures:** 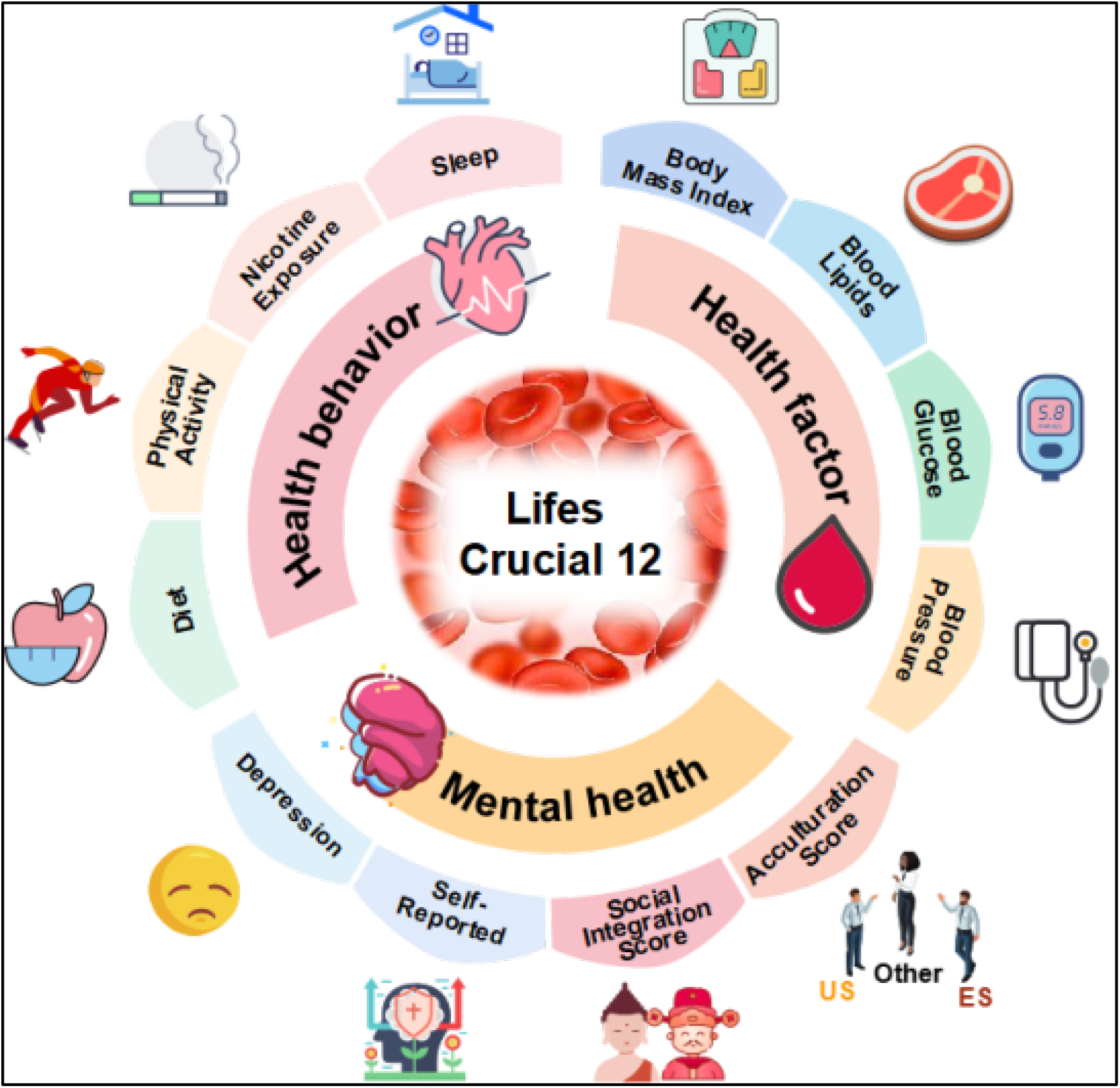

## 1. Introduction

Stroke, the third most common cause of death and disability worldwide, is a severe cardiovascular condition (CVD)(1). It comprises ischemic stroke (commonly known as cerebral infarction) and hemorrhagic stroke (such as parenchymal hemorrhage, ventricular hemorrhage, and subarachnoid hemorrhage), a disease causing the necrosis of brain cells and tissues. The global economic impact of stroke is estimated to surpass $721 billion (0.66% of global GDP)(1). From 1990 to 2019, there has been a significant rise in the disease burden (70.0% increase in stroke occurrence, 43.0% increase in stroke fatality, 102.0% stroke prevalence, and 143.0% DALY prevalence)(1). While the exact cause of stroke remains unclear, noteworthy environmental factors are increasingly acknowledged as crucial factors in stroke development.

stroke is gradually being evaluated as a separate outcome rather than being included in studies as part of a composite CVD outcome(2). It is well known that stroke is strongly associated with poor health behaviors and factors(3). Studies suggest that most CVD events, including stroke, can be prevented by optimizing some of the key indicators of CVH(4). Non-modifiable (e.g., age, gender, race or ethnicity, and genetics)(5-6) and modifiable (e.g., sleep quality, physical activity, glycemic abnormalities, diet, body mass index (BMI), smoking, dyslipidemia, and hypertension) risk factors have been extensively studied in programs for primary and secondary cardiovascular (stroke-oriented) prevention(3, 7-8). Maintaining CVH levels can help decrease the risk of developing CVD, such as smoking cessation, increased physical activity, increased sleep time, and a balanced diet(9-11). The American Heart Association developed the risk factor-based CVH model for the prevention of CVD and stroke in 2010(12). The CVH is measured using 7 traditional health metrics: smoking, body mass index (BMI), physical activity (PA), dietary patterns, total cholesterol, blood pressure (BP), and fasting blood glucose, called Life’s Simple 7 (LS7)(12). Population prevalence of ideal, moderate, or poor levels of each indicator, as well as a total CVH score based on all 7 indicators are associated with CVD(8, 12). The relationship between the LS7 score and stroke outcome has been widely studied in various research papers(4, 8, 13). Despite the usefulness of the LS7 CVH score, limitations have been noted, as it may not be effective in detecting differences within or between individuals. To address this issue, the American Heart Association (AHA) introduced an updated CVH assessment tool called Life’s Essential 8 (LE8) in 2022. This new tool includes considerations for sleep duration and expands the age range covered throughout one’s life(14). The scoring system for each component in LE8 has been revised to range from 0 to 100 points(14). Both the LS7 and LE8 play significant roles in evaluating and preventing the risk of stroke(3-4, 8). In particular, a rise of 1 score in the LS7 rating is linked with an 8% decline in stroke risk, and even slight variations in the score can lower the chances of stroke(15). Moreover, a decrease in the LE8 rating corresponds to an increase in adverse heart-related incidents such as heart disease, heart attacks, strokes, and heart failures(16). Additionally, a study conducted in Kailuan cohort, China, revealed that upholding the top 25% of LE8 scores could prevent half of all cases of atherosclerotic heart disease(17). Additionally, it was observed that an increase in LE8 score to the top tertile led to a 44% decrease in the likelihood of atherosclerotic cardiovascular disease development, along with a 43% decrease in the lifetime risk of disease occurrence(17). Of greater significance, LE8 scores correlate with the status of arterial stiffness (an early pathological indicator of stroke)(18), and enhancement in any LE8 element can potentially lower the risks associated with stroke and arterial stiffness(19).

Recently, cardiovascular medicine specialists have suggested the incorporation of psychological health evaluations in CVH assessment(4). Common risk factors for CVD include depression, anxiety, insomnia, and stress(20-22). This indicates the complexity of psychological health and the challenge of measuring it accurately(4, 14). Psychological well-being includes a variety of aspects (depression, anxiety, acculturation status, suicidal ideation, and social integration), all of which are linked independently to CVD risk and consequences(4). There has been limited research on the psychological aspects of stroke in the past, with a scarcity of available studies. Furthermore, effective emotion regulation is essential for overall emotional and social resilience. It is imperative to develop comprehensive cardiovascular care models that address the management of psychological health.

This study planned to construct a new model of integrated cardiovascular care in NHANES (Life’s Crucial 12, LC12). Due to the difficulty in obtaining the suicide awareness variable, the high number of missing values for the anxiety and perceived stress variables, and the variable for the content of participation in religious activities (used in the assessment of the Social Integration Index) were present in only some of the survey cycles (2005-2006, 2007-2008, year cycle). Therefore, we used the limited data to construct a new comprehensive cardiovascular care model (LC12). The LC12 adds Psychological Health to the LE8 score and consists of 4 Health Behaviors (Diet, Physical Activity, Nicotine Exposure, and Sleep Duration), 4 Health Factors (Body Mass Index, Non-High-Density Lipoprotein Cholesterol, Blood Glucose, and Blood Pressure), and 4 Psychological Health (Depression, Self-Reported Mental Health. Social Integration Score (SNI), and Cultural Adaptation Score), with the full elaboration of LC12 detailed in the Method section.

Depression plays a crucial role in psychological health, as individuals experiencing depression or stress have a higher likelihood of experiencing an ischemic stroke. Pre-stroke depression and distress are closely linked to the severity of a stroke(22). A significant case-control study conducted across multiple centers revealed that depression was connected to a heightened risk of all types of strokes, particularly ischemic strokes(23). Furthermore, enhancements in the Social Network Index (SNI) such as marital status, engagement in religious activities, and having a larger social circle can impact an individual’s mental health positively(24). For example, social networks help individuals achieve better status in terms of emotional aspects (e.g., detachment, love, empathy,) instrumental aspects (e.g., travel, help with household chores), and informational aspects (e.g., health education, increased physical activity, and timely access to medical care)(25-26). Furthermore, membership in a religious group with health beliefs(27) and social network status, characterized by the quality and quantity of personal and community relationships, can influence health behaviors and outcomes(25), e.g., smaller social networks and a lack of social support are associated with an increased incidence of stroke and myocardial infarction, a higher prevalence of dementia(28-29), and an increased risk of poststroke depression and death(30). An increased risk of stroke due to poor SNI has also been observed in women with myocardial ischemia(31). Comprehending the social connections of individuals and their impact on health risks and outcomes could offer important perspectives for modeling human health and identifying potential treatment options. Research has indicated that individuals who have experienced a stroke may have an increased likelihood of reporting mental health issues on certain days(32). Severe mental disorders can lead to physiological disruptions, exacerbating pre-existing health conditions and notably raising the occurrence of strokes(33-35). Studies have shown that acculturation could be significantly linked to whether the native language is spoken in the community, the connection between birthplace and current living location, and the length of time residing in the current region(36-37). Cognitive impairment evaluated by the MMSE is impacted by both acculturation and structural assimilation, with results also affected by the interviewer’s language(38). Current research lacks information on acculturation in stroke management. Furthermore, individuals with lower levels of acculturation, regardless of language proficiency, experience inferior recovery post-stroke(37).

LS7 and LE8 are recognized risk factors for stroke, however, the crucial role of Psychological Health in stroke development was overlooked. The aforementioned studies lay the groundwork for stroke prevention and the creation of the LC12 score model discussed in this research. Consequently, we conducted a cross-sectional study using data from the 2005-2008 NHANES to develop a more comprehensive integrated cardiovascular care model (LC12) that considers Health Behavior, Health Factors, and Psychological Health. Furthermore, we delved into the relationship between LC12 and the risk of stroke.

## 2. METHODS

### 2.1 Study Population

NHANES is designed to be a nationally representative sample of the U.S. population. Participants are selected from the noninstitutionalized U.S. civilian population using a complex, stratified, multistage probability cluster sampling design. The survey was conducted periodically through 1999 and continuously thereafter. It is a cross-sectional survey designed to assess the health and nutritional status of adults and children in the United States. The survey protocol was approved by the National Center for Health Statistics (NCHS) Research Ethics Review Board and informed consent was obtained from each participant. Although the U.S. NHANES publishes data through 2020, information on our study’s process variable participation in religious activities was included in only two survey cycles (2005-2006, 2007-2008), so we selected only data collected in 2005-2008. The raw data for this study consisted of individuals aged ≥ 20 years and older who were not pregnant, and this study ultimately included 4,478 participants, which, by assigning population weights, is representative of the general population of 94,054,134 individuals in the U.S. The specific population screening process is shown in **Figure 1**.

**Figure 1.**
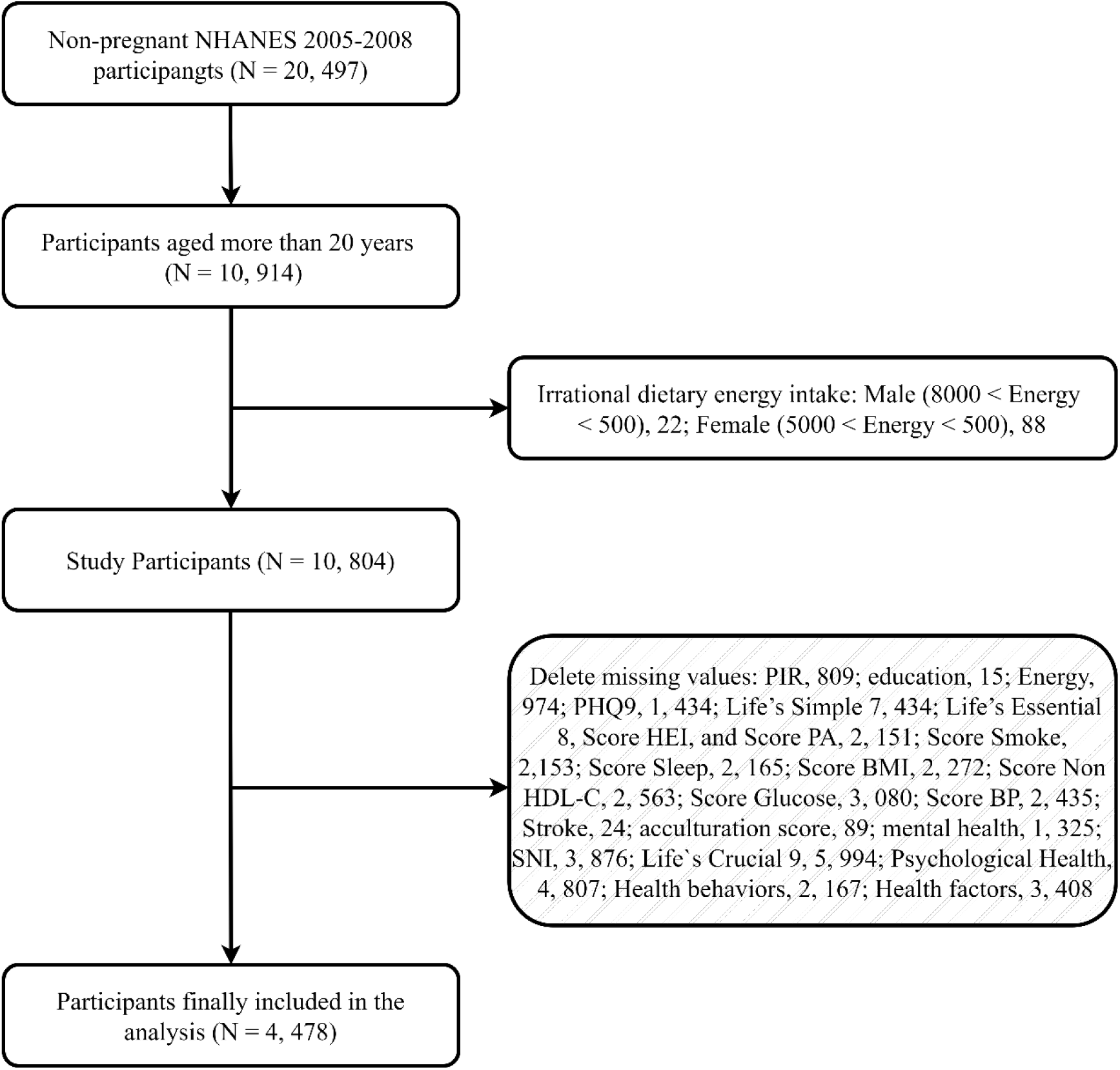
Flowchart of the exclusion process used to derive the healthy reference population from NHANES (2005-2008).

### 2.2 Life’s Crucial 12 Construction

We propose to consider updating the indicators for Psychological Health(4) from the original eight indicators of LE8 to construct a comprehensive cardiovascular care model (Life’s Crucial 12, LC12) to more fully account for inter-individual differences and intra-individual variability. The aim is for researchers, health systems, and policy makers to create standardized tools to measure and monitor CVH in individuals and populations. The evaluation criteria and detailed calculations for the LE8 have been reported in previous studies(14), following the model of the LE8 score for the 4 health behaviors (diet, physical activity, nicotine exposure, and sleep duration), the 4 health factors (body mass index, non-high-density lipoprotein (NHLDL) cholesterol, glucose, and blood pressure), and their scoring criteria(14). Sub-questionnaires for the LC12 scores included four health behaviors (diet, physical activity, nicotine exposure, and sleep duration), four health factors (body mass index, non-HDL cholesterol, blood glucose, and blood pressure), and four psychological health (Depression(39), Self-reported mental health, , Social Network Index (SNI)(24), Acculturation score(40)). The Psychological Health score is the arithmetic mean of the above four evaluation indicators, and the LC12 score is the arithmetic mean of 12 evaluation indicators in the three domains of Health Behavior. The LC12 score is the arithmetic mean of 12 indicators in the three domains of health behavior, health factors, and mental health. In short, the 12 CVH indicators range from 0 to 100 points, and the total LC12 score is calculated as the arithmetic mean of the 12 indicators. The LC12 scores are categorized into low (0 to 49 score), medium (50 to 79 score), and high (80 - 100 score) CVH. as shown in **Table 1**. The evaluation criteria as well as the calculation of the scores have been fully described in the **Supplementary Material (Method section)** regarding the construct of psychological health.

**Table 1.**
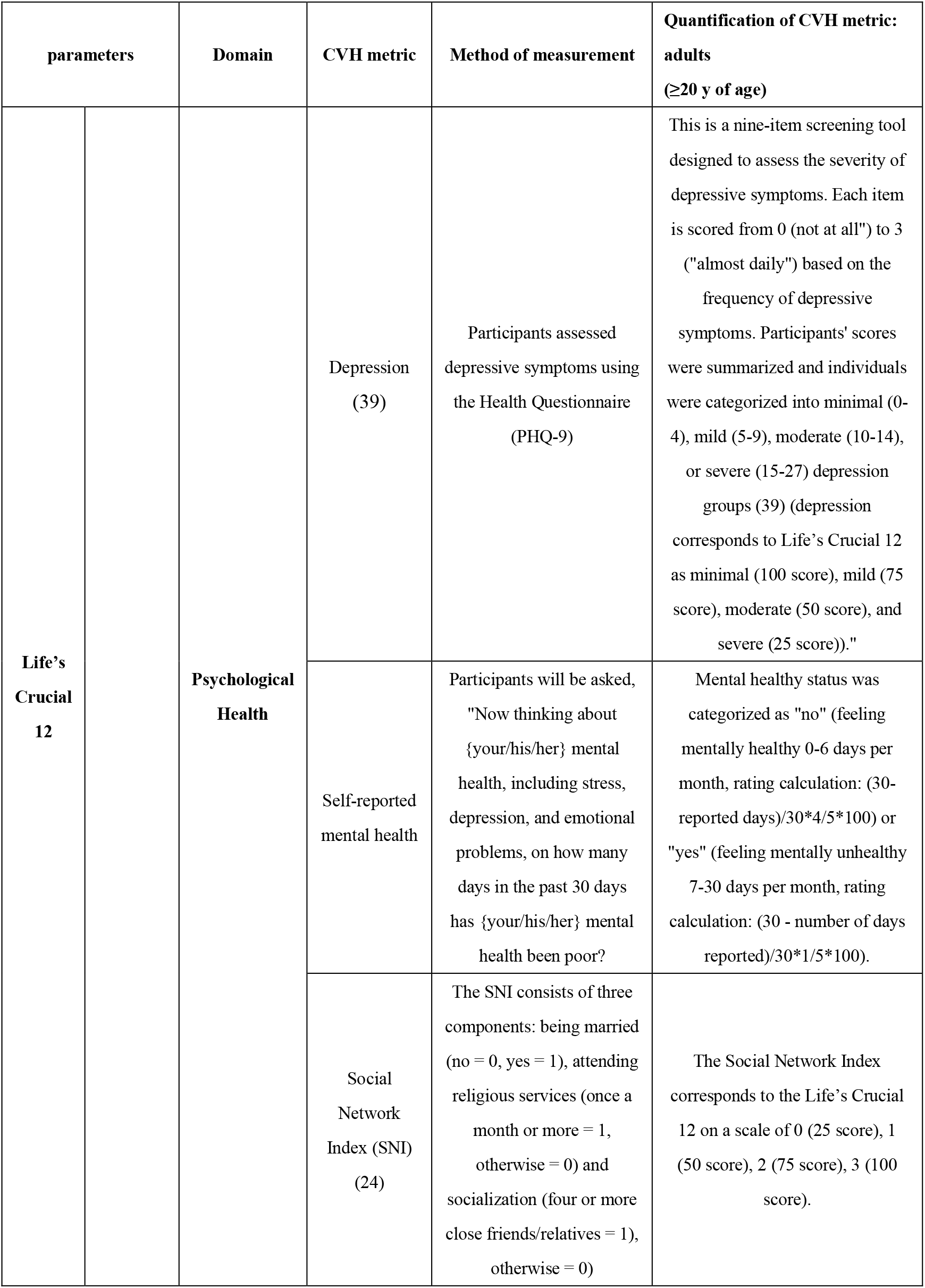
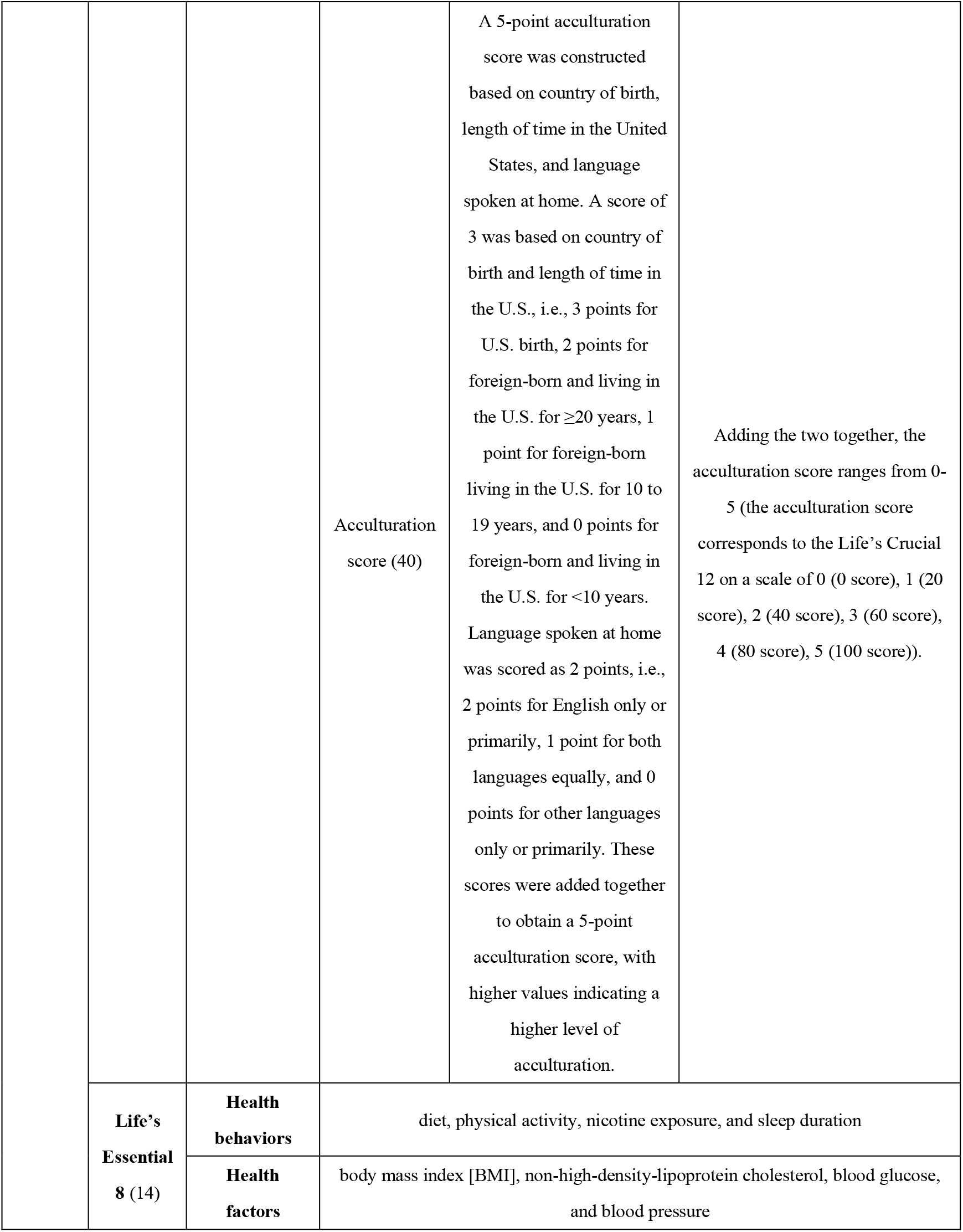
Indicators for CVH measurement and quantitative assessment.

| parameters |  | Domain | CVH metric | Method of measurement | Quantification of CVH metric:<br>adults<br>(≥20 y of age) |
| --- | --- | --- | --- | --- | --- |
| Life's<br>Crucial<br>12 |  | Psychological<br>Health | Depression<br>(39) | Participants assessed depressive symptoms using the Health Questionnaire (PHQ-9) | This is a nine-item screening tool designed to assess the severity of depressive symptoms. Each item is scored from 0 (not at all") to 3 ("almost daily") based on the frequency of depressive symptoms. Participants' scores were summarized and individuals were categorized into minimal (0-4), mild (5-9), moderate (10-14), or severe (15-27) depression groups (39) (depression corresponds to Life's Crucial 12 as minimal (100 score), mild (75 score), moderate (50 score), and severe (25 score))." |
|  |  |  | Self-reported mental health | Participants will be asked, "Now thinking about {your/his/her} mental health, including stress, depression, and emotional problems, on how many days in the past 30 days has {your/his/her} mental health been poor?" | Mental healthy status was categorized as "no" (feeling mentally healthy 0-6 days per month, rating calculation: (30-reported days)/30*4/5*100) or "yes" (feeling mentally unhealthy 7-30 days per month, rating calculation: (30 - number of days reported)/30*1/5*100). |
|  |  |  | Social Network Index (SNI)<br>(24) | The SNI consists of three components: being married (no = 0, yes = 1), attending religious services (once a month or more = 1, otherwise = 0) and socialization (four or more close friends/relatives = 1), otherwise = 0) | The Social Network Index corresponds to the Life's Crucial 12 on a scale of 0 (25 score), 1 (50 score), 2 (75 score), 3 (100 score). |
|  |  |  | Acculturation score (40) | A 5-point acculturation score was constructed based on country of birth, length of time in the United States, and language spoken at home. A score of 3 was based on country of birth and length of time in the U.S., i.e., 3 points for U.S. birth, 2 points for foreign-born and living in the U.S. for $\geq 20$ years, 1 point for foreign-born living in the U.S. for 10 to 19 years, and 0 points for foreign-born and living in the U.S. for $< 10$ years. Language spoken at home was scored as 2 points, i.e., 2 points for English only or primarily, 1 point for both languages equally, and 0 points for other languages only or primarily. These scores were added together to obtain a 5-point acculturation score, with higher values indicating a higher level of acculturation. | Adding the two together, the acculturation score ranges from 0-5 (the acculturation score corresponds to the Life's Crucial 12 on a scale of 0 (0 score), 1 (20 score), 2 (40 score), 3 (60 score), 4 (80 score), 5 (100 score)). |
|  | **Life's Essential 8 (14)** | **Health behaviors** | diet, physical activity, nicotine exposure, and sleep duration |  |  |
|  |  | **Health factors** | body mass index [BMI], non-high-density-lipoprotein cholesterol, blood glucose, and blood pressure |  |  |

### 2.3 Assessment of stroke and Other Variables of Interest

We defined stroke based on self-reported interview data obtained from the Medical Conditions Questionnaire. Participants were asked the following question, “Has a doctor or other health professional ever told you that you had a stroke?”, If they answered yes, they were categorized in the stroke group. Conversely, participants who denied being told they had a stroke were included in the non-stroke group(41). Life’s Simple 7 (LS7) framework encompasses various factors that contribute to overall health, including physical activity, smoking habits, BMI, dietary patterns, blood glucose levels, blood pressure, and total cholesterol levels(13). Demographic information was collected by questionnaire, including age, sex, race (non-Hispanic white people, non-Hispanic black people, Hispanic people, and others), family income to poverty ratio (less than 1.30, 1.30 to <3.00, 3.00 to <5.00, 5.00 and above, represents the ratio of family income to the federal poverty threshold, adjusting for household size. A higher ratio indicates a higher levels of income), education (less than high school (Less than 11th grade), high school graduate/general education, some college, or above (College graduate or above)).

### 2.4 Statistical Analysis

All remaining statistical analyses were performed using R software (4.2.2, https://cran.r-project.org/bin/windows/base/old/4.2.2/), with statistical tests being two-sided and considered statistically significant when the P-value < 0.05. To obtain statistics representative of U.S. adults, we utilized oversampling, stratification, and clustering techniques in the NHANES. We weighted the NHANES data for analysis. Baseline population characteristics were reported for continuous variables and as numerical values (percentage) for categorical variables. Demographic characteristics of participants stroke status were assessed using weighted chi-squared test was used to assess disparities in categorical variables. Weighted Student’s t-test or weighted linear regression was employed to analyze differences among continuous variables. Life’s Crucial 12 and Life’s Essential 8 scores are categorized as Low (0 to 49), Medium (50 to 79), and High (80 - 100), with Low (0 to 49) as the reference, Therefore, the total score for Life’s Simple 7 has been categorized as Poor (0 to 7), Intermediate (8 to 10), and Ideal (11 to 14)(13), with Inadequate (0 to 7 points) as a reference. The odds ratios (OR) with 95% confidence intervals (CI) for the association between Life`s Crucial 9, Psychological Health, Health behaviors, Health factors, and stroke risk were estimated through applied univariate logistic regression and multivariate logistic regression models. Model 1: Did not adjust any covariates. Model 2: Adjusted for age, gender, race. Model 3: Adjusted for age, gender, race, PIR (family income to poverty ratio), education. Restricted cubic spline (RCS) plots are used to show trends for variables that are significant in the logistic regression portion. RCS plots are used to determine whether there is a nonlinear association between exposure factors Life’s Crucial 12, Psychological Health, Health behaviors, Health factors, and stroke risk.

### 2.5 Sensitivity analysis

Meanwhile, we performed sensitivity analyses: (1) The simple division of race into white people and black people. (2) Age as a categorical variable: age (20–44, 45–64, 65, and older). (3) Multivariate logistic regression was used to analyze the association between Life’s Simple 7 and stroke risk (Psychological health and sleep not taken into account). (4) Multivariate logistic regression was used to analyze the association between Life’s Essential 8 and stroke risk (Psychological health not taken into account).

## 3. Result

### 3.1 Characteristics of participants

The baseline characteristics of the study population are summarized in **Table 2** according to stroke. Among the 4, 478 participants aged 20 years or older who were included (mean age 56.568 years), 250 participants had stroke (mean age 66.070 years). Compared to Non-Stroke participants, stroke had lower CVH (Life’s Simple 7, Life’s Essential 8, Life’s Crucial 12) scores, which were 6.121, 54.113, 59.060. Importantly, patients with stroke had lower scores for Psychological Health, Health behaviors, Health factors, 68.953, 52.775, 55.451 respectively. Overall, Energy, Life ‘s Crucial 12, Psychological Health, Health behaviors, Health factors, Life’s Simple 7, Life’s Essential 8. Score PA, Score Sleep, Score BMI, Score Glucose, Mental Health, SNI, Age, Sex, and PIR had statistically significant differences between Non-Stroke and Stroke participants.

**Table 2.**
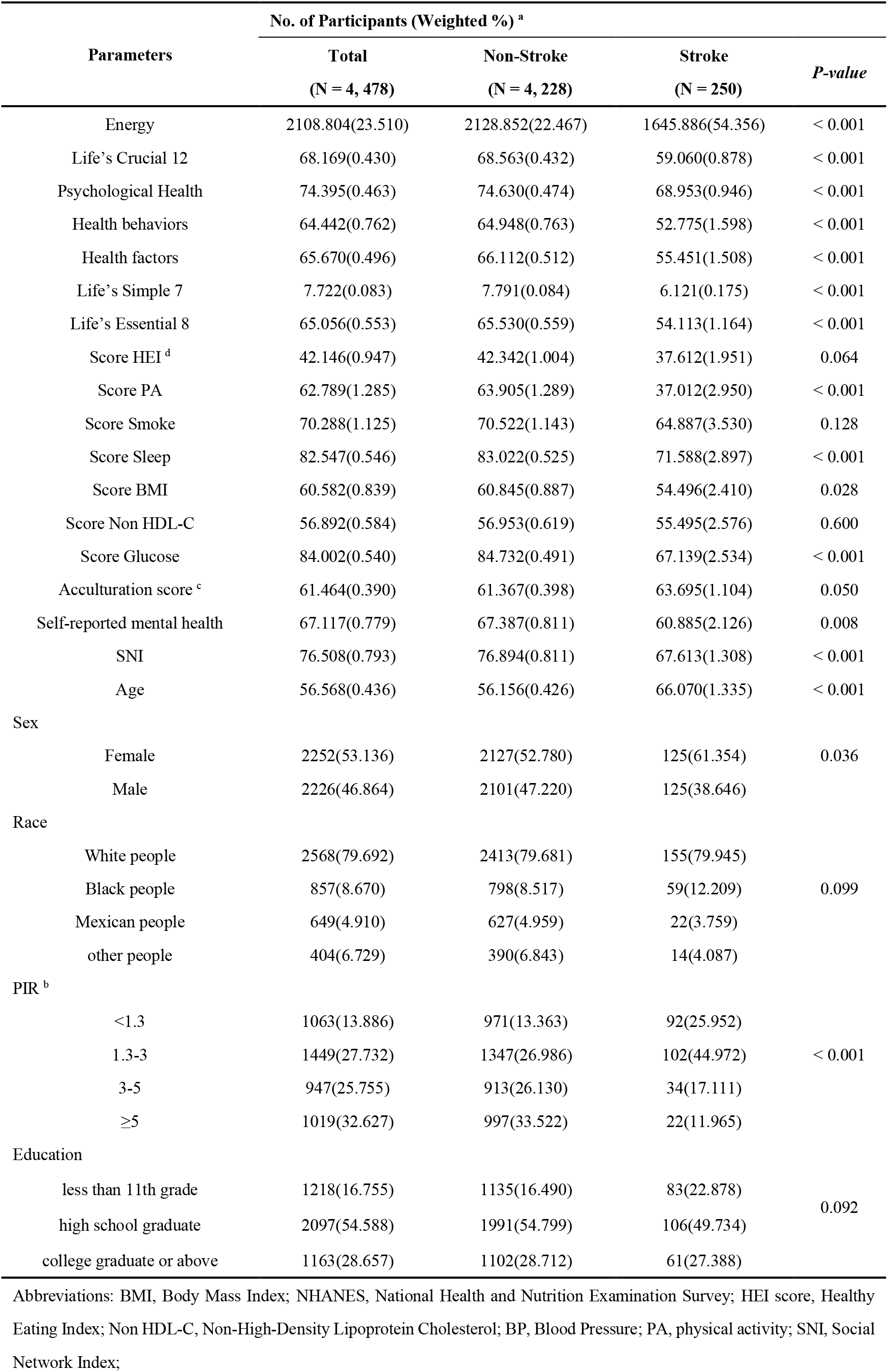
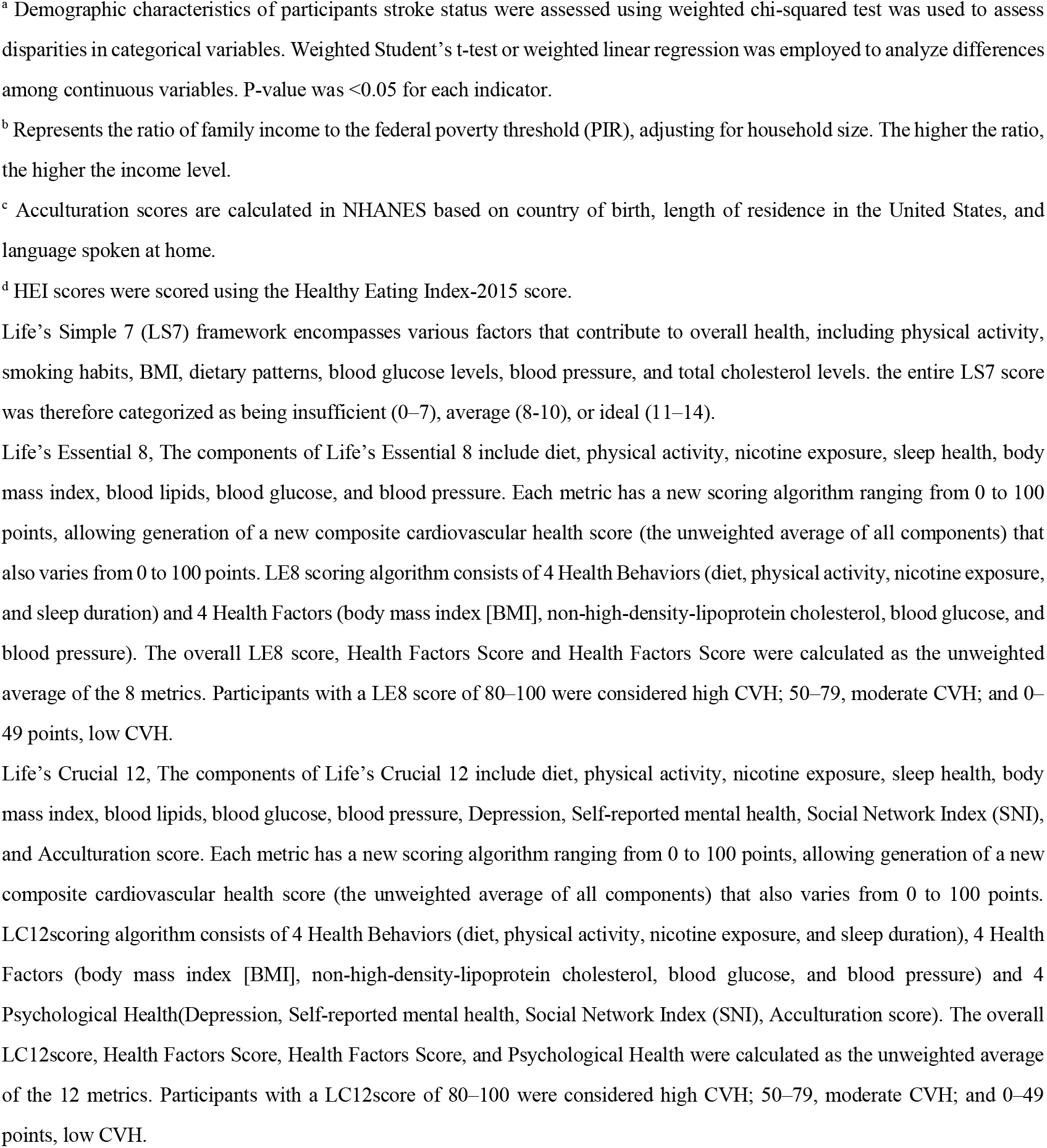
Socio-demographic characteristics of America adults with Stroke by NHANES survey cycle, 2005-2008.

| Parameters | No. of Participants (Weighted %) <sup>a</sup> |  |  | *P-value* |
| --- | --- | --- | --- | --- |
|  | Total<br>(N = 4, 478) | Non-Stroke<br>(N = 4, 228) | Stroke<br>(N = 250) |  |
| Energy | 2108.804(23.510) | 2128.852(22.467) | 1645.886(54.356) | < 0.001 |
| Life's Crucial 12 | 68.169(0.430) | 68.563(0.432) | 59.060(0.878) | < 0.001 |
| Psychological Health | 74.395(0.463) | 74.630(0.474) | 68.953(0.946) | < 0.001 |
| Health behaviors | 64.442(0.762) | 64.948(0.763) | 52.775(1.598) | < 0.001 |
| Health factors | 65.670(0.496) | 66.112(0.512) | 55.451(1.508) | < 0.001 |
| Life's Simple 7 | 7.722(0.083) | 7.791(0.084) | 6.121(0.175) | < 0.001 |
| Life's Essential 8 | 65.056(0.553) | 65.530(0.559) | 54.113(1.164) | < 0.001 |
| Score HEI <sup>d</sup> | 42.146(0.947) | 42.342(1.004) | 37.612(1.951) | 0.064 |
| Score PA | 62.789(1.285) | 63.905(1.289) | 37.012(2.950) | < 0.001 |
| Score Smoke | 70.288(1.125) | 70.522(1.143) | 64.887(3.530) | 0.128 |
| Score Sleep | 82.547(0.546) | 83.022(0.525) | 71.588(2.897) | < 0.001 |
| Score BMI | 60.582(0.839) | 60.845(0.887) | 54.496(2.410) | 0.028 |
| Score Non HDL-C | 56.892(0.584) | 56.953(0.619) | 55.495(2.576) | 0.600 |
| Score Glucose | 84.002(0.540) | 84.732(0.491) | 67.139(2.534) | < 0.001 |
| Acculturation score <sup>c</sup> | 61.464(0.390) | 61.367(0.398) | 63.695(1.104) | 0.050 |
| Self-reported mental health | 67.117(0.779) | 67.387(0.811) | 60.885(2.126) | 0.008 |
| SNI | 76.508(0.793) | 76.894(0.811) | 67.613(1.308) | < 0.001 |
| Age | 56.568(0.436) | 56.156(0.426) | 66.070(1.335) | < 0.001 |
| Sex |  |  |  |  |
| Female | 2252(53.136) | 2127(52.780) | 125(61.354) | 0.036 |
| Male | 2226(46.864) | 2101(47.220) | 125(38.646) |  |
| Race |  |  |  |  |
| White people | 2568(79.692) | 2413(79.681) | 155(79.945) | 0.099 |
| Black people | 857(8.670) | 798(8.517) | 59(12.209) |  |
| Mexican people | 649(4.910) | 627(4.959) | 22(3.759) |  |
| other people | 404(6.729) | 390(6.843) | 14(4.087) |  |
| PIR <sup>b</sup> |  |  |  |  |
| <1.3 | 1063(13.886) | 971(13.363) | 92(25.952) | < 0.001 |
| 1.3-3 | 1449(27.732) | 1347(26.986) | 102(44.972) |  |
| 3-5 | 947(25.755) | 913(26.130) | 34(17.111) |  |
| ≥5 | 1019(32.627) | 997(33.522) | 22(11.965) |  |
| Education |  |  |  |  |
| less than 11th grade | 1218(16.755) | 1135(16.490) | 83(22.878) | 0.092 |
| high school graduate | 2097(54.588) | 1991(54.799) | 106(49.734) |  |
| college graduate or above | 1163(28.657) | 1102(28.712) | 61(27.388) |  |

Abbreviations: BMI, Body Mass Index; NHANES, National Health and Nutrition Examination Survey; HEI score, Healthy Eating Index; Non HDL-C, Non-High-Density Lipoprotein Cholesterol; BP, Blood Pressure; PA, physical activity; SNI, Social Network Index;
<sup>a</sup> Demographic characteristics of participants stroke status were assessed using weighted chi-squared test was used to assess disparities in categorical variables. Weighted Student's t-test or weighted linear regression was employed to analyze differences among continuous variables. P-value was <0.05 for each indicator.
<sup>b</sup> Represents the ratio of family income to the federal poverty threshold (PIR), adjusting for household size. The higher the ratio, the higher the income level.
<sup>c</sup> Acculturation scores are calculated in NHANES based on country of birth, length of residence in the United States, and language spoken at home.
<sup>d</sup> HEI scores were scored using the Healthy Eating Index-2015 score.
Life's Simple 7 (LS7) framework encompasses various factors that contribute to overall health, including physical activity, smoking habits, BMI, dietary patterns, blood glucose levels, blood pressure, and total cholesterol levels. the entire LS7 score was therefore categorized as being insufficient (0–7), average (8–10), or ideal (11–14).
Life's Essential 8, The components of Life's Essential 8 include diet, physical activity, nicotine exposure, sleep health, body mass index, blood lipids, blood glucose, and blood pressure. Each metric has a new scoring algorithm ranging from 0 to 100 points, allowing generation of a new composite cardiovascular health score (the unweighted average of all components) that also varies from 0 to 100 points. LE8 scoring algorithm consists of 4 Health Behaviors (diet, physical activity, nicotine exposure, and sleep duration) and 4 Health Factors (body mass index [BMI], non-high-density-lipoprotein cholesterol, blood glucose, and blood pressure). The overall LE8 score, Health Factors Score and Health Factors Score were calculated as the unweighted average of the 8 metrics. Participants with a LE8 score of 80–100 were considered high CVH; 50–79, moderate CVH; and 0–49 points, low CVH.
Life's Crucial 12, The components of Life's Crucial 12 include diet, physical activity, nicotine exposure, sleep health, body mass index, blood lipids, blood glucose, blood pressure, Depression, Self-reported mental health, Social Network Index (SNI), and Acculturation score. Each metric has a new scoring algorithm ranging from 0 to 100 points, allowing generation of a new composite cardiovascular health score (the unweighted average of all components) that also varies from 0 to 100 points. LC12scoring algorithm consists of 4 Health Behaviors (diet, physical activity, nicotine exposure, and sleep duration), 4 Health Factors (body mass index [BMI], non-high-density-lipoprotein cholesterol, blood glucose, and blood pressure) and 4 Psychological Health (Depression, Self-reported mental health, Social Network Index (SNI), Acculturation score). The overall LC12score, Health Factors Score, Health Factors Score, and Psychological Health were calculated as the unweighted average of the 12 metrics. Participants with a LC12score of 80–100 were considered high CVH; 50–79, moderate CVH; and 0–49 points, low CVH.

### 3.2 Univariate logistic regression models of Life’s Crucial 12 with stroke for participants

In Model 1, compared with the Low group of LC 12 participants, the OR (95% CI) was 0.215 (0.135, 0.340), 0.034 (0.015, 0.081) for the LC 12 Moderate and Height groups; the OR (95% CI) for the Psychological Health Moderate and Height groups was 0.407 (0.257, 0.644), 0.199(0.112, 0.352), OR (95% CI) for Health Behaviors moderate and high groups was 0.523(0.383, 0.715), 0.180(0.104, 0.313), OR (95% CI) for Health Factors moderate and high groups was 0.95%. CI) was 0.389 (0.287, 0.525), 0.211 (0.117, 0.380). In Model 2, compared with the Low group of LC 12 participants, the OR (95% CI) was 0.166 (0.101, 0.272), 0.030 (0.013, 0.073) for the LC 12 Moderate and High groups, 0.274 (0.169, 0.443), 0.138 (0.082, 0.231) for the Psychological Health Moderate and High groups, and 0.274 (0.169, 0.443), 0.138 (0.082, 0.231), OR (95% CI) for Health Behaviors moderate and high groups was 0.505 (0.375, 0.680), 0.161 (0.095, 0.274), OR (95% CI) for Health Factors moderate and high groups was 0.505 (0.375, 0.680), 0.161 (0.095, 0.274), OR (95% CI) for Health Factors moderate and high groups was 0.030 (0.013, 0.073). CI) was 0.428 (0.311,0.589), 0.308 (0.167, 0.569). In Model 3, compared with the Low group of LC 12 participants, the OR (95% CI) was 0.198 (0.118, 0.331), 0.042 (0.016, 0.105) for the LC 12 Moderate and High groups, 0.335 for the Psychological Health Moderate and High groups 0.335 (0.206, 0.544), 0.183(0.101, 0.331), OR (95% CI) for Health Behaviors moderate and high groups was 0.554 (0.401, 0.766), 0.190 (0.114, 0.315), OR (95% CI) for Health Factors moderate and high groups was 0.95%. CI) was 0.456 (0.330, 0.629), 0.340 (0.182, 0.634). Overall, LC 12, Psychological Health, Health Behaviors, and Health Factors reduced the risk of Stroke, possibly a protective factor for Stroke (*P-value* < 0.001), and there was a significant between-group trend (*P-trend* < 0.001). As shown in **Table 3**.

**Table 3.** **U**nivariate logistic regression models of Life’s Crucial 12 with stroke for participants.

| Parameter | model 1 |  | Model 2 |  | Model 3 |  |
| --- | --- | --- | --- | --- | --- | --- |
|  | OR (95%CI) | P-value | OR (95%CI) | P-value | OR (95%CI) | P-value |
| **Life's Crucial 12 <sup>a</sup>** |  |  |  |  |  |  |
| Low | ref |  | ref |  | ref |  |
| Moderate | 0.215(0.135,0.340) | <0.001 | 0.166(0.101,0.272) | <0.001 | 0.198(0.118,0.331) | <0.001 |
| High | 0.034(0.015,0.081) | <0.001 | 0.030(0.013,0.073) | <0.001 | 0.042(0.016,0.105) | <0.001 |
| *P-trend* | <0.001 |  | <0.001 |  | <0.001 |  |
| **Psychological Health <sup>a</sup>** |  |  |  |  |  |  |
| Low | ref |  | ref |  | ref |  |
| Moderate | 0.407(0.257,0.644) | <0.001 | 0.274(0.169,0.443) | <0.001 | 0.335(0.206,0.544) | <0.001 |
| High | 0.199(0.112,0.352) | <0.001 | 0.138(0.082,0.231) | <0.001 | 0.183(0.101,0.331) | <0.001 |
| *P-trend* | <0.001 |  | <0.001 |  | <0.001 |  |
| **Health Behaviors <sup>a</sup>** |  |  |  |  |  |  |
| Low | ref |  | ref |  | ref |  |
| Moderate | 0.523(0.383,0.715) | <0.001 | 0.505(0.375,0.680) | <0.001 | 0.554(0.401,0.766) | <0.001 |
| High | 0.180(0.104,0.313) | <0.001 | 0.161(0.095,0.274) | <0.001 | 0.190(0.114,0.315) | <0.001 |
| *P-trend* | <0.001 |  | <0.001 |  | <0.001 |  |
| **Health Factors <sup>a</sup>** |  |  |  |  |  |  |
| Low | ref |  | ref |  | ref |  |
| Moderate | 0.389(0.287,0.525) | <0.001 | 0.428(0.311,0.589) | <0.001 | 0.456(0.330,0.629) | <0.001 |
| High | 0.211(0.117,0.380) | <0.001 | 0.308(0.167,0.569) | <0.001 | 0.340(0.182,0.634) | 0.002 |
| *P-trend* | <0.001 |  | <0.001 |  | <0.001 |  |

Model 1: Did not adjust any covariates.
Model 2: Adjusted for age, gender, race.
Model 3: Adjusted for age, gender, race, PIR (family income to poverty ratio), education.
<sup>a</sup> Categorical variable.
Life's Crucial 12, The components of Life's Crucial 12 include diet, physical activity, nicotine exposure, sleep health, body mass index, blood lipids, blood glucose, blood pressure, Depression, Self-reported mental health, Social Network Index (SNI), and Acculturation score. Each metric has a new scoring algorithm ranging from 0 to 100 points, allowing generation of a new composite cardiovascular health score (the unweighted average of all components) that also varies from 0 to 100 points. LC12scoring algorithm consists of 4 Health Behaviors (diet, physical activity, nicotine exposure, and sleep duration), 4 Health Factors (body mass index [BMI], non-high-density-lipoprotein cholesterol, blood glucose, and blood pressure) and 4 Psychological Health (Depression, Self-reported mental health, Social Network Index (SNI), Acculturation score). The overall LC12score, Health Factors Score, Health Factors Score, and Psychological Health were calculated as the unweighted average of the 12 metrics. Participants with a LC12score of 80–100 were considered high CVH; 50–79, moderate CVH; and 0–49 points, low CVH.

### 3.3 Multiple logistic regression models of Life’s Crucial 12 with stroke for participants

In Model 3, the ORs for the LC 12 Moderate and High groups compared to the Low group of LC 12 participants were 0.431 (0.226, 0.822), 0.212 (0.060, 0.755), the ORs (95% CI) for the Psychological Health Moderate and High groups were 0.536 ( 0.297, 0.967), 0.357(0.178, 0.713), OR (95% CI) for Health Behaviors moderate and high groups was 0.759 (0.552, 1.043), 0.334 (0.179, 0.623), OR (95% CI) for Health Factors moderate and high groups was 0.759 (0.552, 1.043), 0.334 (0.179, 0.623), and OR (95% CI) for Health Factors moderate and high groups was 0.212 (0.060, 0.755). CI) were 0.565 (0.406, 0.786), 0.533 (0.286, 0.994). In Model 3, Life’s Crucial 12, Psychological Health, Health behaviors, and Health factors as continuous variables in Model 3 still reduces the risk of Stroke (results were: 0.926 (0.909, 0.942), 0.966 (0.955, 0.977), 0.969 (0.961, 0.978), and 0.975 (0.963, 0.987)), which are protective factors for Stroke. In Model 3, Life’s Crucial 12, Psychological Health, Health behaviors, Health factors as continuous variables still reduced the risk of developing Stroke (results were: 0.926 (0.909, 0.942), 0.966 (0.955, 0.977), 0.969 (0.961, 0.978), 0.975 (0.963, 0.987)), which are protective factors for Stroke. LC12, Psychological health perse, Health behaviors perse, and Health factors perse as continuous variables still reduced the risk of Stroke (results: 0.994 (0.993, 0.995), 0.997 (0.996, 0.998), 0.996 (0.996, 0.998), 0.996 (0.995, 0.997), 0.997 (0.995, 0.998)), a protective factor for Stroke.

Overall, LC 12, Psychological Health, Health Behaviors, and Health Factors reduced the risk of Stroke, and may be protective factors for Stroke, and there was a significant between-group trend (*P-trend* < 0.001). As shown in **Table** 4.We found that the OR values of LC 12, Psychological Health, Health behaviors, Health factors scores and stroke dropped sharply in the lower range, and then the ORs remained stable in the higher values, no nonlinear relationship was observed between LC 12 scores and Stroke (*P-Total* < 0.001, *P-Nonlinearity* = 0.271). The minimal threshold for the beneficial association was 66.997 scores (estimate OR = 1). In Psychological Health scores No nonlinear relationship was observed between Health Behaviors scores and Stroke (*P-Total* < 0.001, *P-Nonlinearity* = 0.491). The minimal threshold for the beneficial association was 77.200 scores (estimate OR = 1). A nonlinear relationship was observed (*P-Total* < 0.001, *P-Nonlinearity* = 0.356). The minimal threshold for the beneficial association was 63.386 scores (estimate OR = 1). No nonlinear relationship was observed between Health Factors scores and Stroke (*P-Total* < 0.001, *P-Nonlinearity* = 0.065). The minimal threshold for the beneficial association was 64.673 scores (estimate OR = 1). As shown in **Figure 2**.

**Table 4.** Multiple logistic regression models of Life’s Crucial 12 with stroke for participants.

| Parameter | model 1 |  | Model 2 |  | Model 3 |  |
| --- | --- | --- | --- | --- | --- | --- |
|  | OR (95%CI) | P-value | OR (95%CI) | P-value | OR (95%CI) | P-value |
| Life's Crucial 12 <sup>a</sup> | 0.928(0.915,0.941) | <0.001 | 0.922(0.906,0.938) | <0.001 | 0.926(0.909,0.942) | <0.001 |
| Life's Crucial 12 perse <sup>a</sup> | 0.994(0.993,0.995) | <0.001 | 0.994(0.993,0.995) | <0.001 | 0.994(0.993,0.995) | <0.001 |
| **Life's Crucial 12 <sup>b</sup>** |  |  |  |  |  |  |
| Low | ref |  | ref |  | ref |  |
| Moderate | 0.542(0.289,1.015) | 0.055 | 0.401(0.208,0.774) | 0.009 | 0.431(0.226,0.822) | 0.015 |
| High | 0.257(0.076,0.867) | 0.03 | 0.180(0.053,0.609) | 0.008 | 0.212(0.060,0.755) | 0.020 |
| *P-trend* | <0.001 |  | <0.001 |  | <0.001 |  |
| Psychological Health <sup>a</sup> | 0.969(0.959,0.980) | <0.001 | 0.960(0.951,0.970) | <0.001 | 0.966(0.955,0.977) | <0.001 |
| Psychological Health perse <sup>a</sup> | 0.997(0.996,0.998) | <0.001 | 0.996(0.995,0.997) | <0.001 | 0.997(0.996,0.998) | <0.001 |
| **Psychological Health <sup>b</sup>** |  |  |  |  |  |  |
| Low | ref |  | ref |  | ref |  |
| Moderate | 0.621(0.366,1.052) | 0.074 | 0.499(0.277,0.898) | 0.023 | 0.536(0.297,0.967) | 0.040 |
| High | 0.388(0.201,0.750) | 0.007 | 0.328(0.173,0.622) | 0.002 | 0.357(0.178,0.713) | 0.007 |
| *P-trend* | <0.001 |  | <0.001 |  | <0.001 |  |
| Health Behaviors <sup>a</sup> | 0.971(0.963,0.979) | <0.001 | 0.967(0.958,0.975) | <0.001 | 0.969(0.961,0.978) | <0.001 |
| Health Behaviors perse <sup>a</sup> | 0.996(0.995,0.997) | <0.001 | 0.995(0.994,0.997) | <0.001 | 0.996(0.995,0.997) | <0.001 |
| **Health Behaviors <sup>b</sup>** |  |  |  |  |  |  |
| Low | ref |  | ref |  | ref |  |
| Moderate | 0.755(0.547,1.042) | 0.084 | 0.743(0.547,1.008) | 0.056 | 0.759(0.552,1.043) | 0.084 |
| High | 0.346(0.185,0.646) | 0.002 | 0.323(0.173,0.602) | 0.001 | 0.334(0.179,0.623) | 0.002 |
| *P-trend* | <0.001 |  | <0.001 |  | <0.001 |  |
| Health Factors <sup>a</sup> | 0.968(0.957,0.978) | <0.001 | 0.973(0.961,0.985) | <0.001 | 0.975(0.963,0.987) | <0.001 |
| Health Factors perse <sup>a</sup> | 0.996(0.994,0.997) | <0.001 | 0.996(0.995,0.998) | <0.001 | 0.997(0.995,0.998) | <0.001 |
| **Health Factors <sup>b</sup>** |  |  |  |  |  |  |
| Low | ref |  | ref |  | ref |  |
| Moderate | 0.487(0.351,0.674) | <0.001 | 0.557(0.404,0.767) | 0.001 | 0.565(0.406,0.786) | 0.003 |
| High | 0.320(0.180,0.569) | <0.001 | 0.532(0.292,0.967) | 0.040 | 0.533(0.286,0.994) | 0.048 |
| *P-trend* | <0.001 |  | <0.001 |  | <0.001 |  |

Model 1: Did not adjust any covariates.
Model 2: Adjusted for age, gender, race.
Model 3: Adjusted for age, gender, race, PIR (family income to poverty ratio), education.
<sup>a</sup> Continuous variable; <sup>b</sup> Categorical variable.
Life's Crucial 12, The components of Life's Crucial 12 include diet, physical activity, nicotine exposure, sleep health, body mass index, blood lipids, blood glucose, blood pressure, Depression, Self-reported mental health, Social Network Index (SNI), and Acculturation score. Each metric has a new scoring algorithm ranging from 0 to 100 points, allowing generation of a new composite cardiovascular health score (the unweighted average of all components) that also varies from 0 to 100 points. LC12scoring algorithm consists of 4 Health Behaviors (diet, physical activity, nicotine exposure, and sleep duration), 4 Health Factors (body mass index [BMI], non-high-density-lipoprotein cholesterol, blood glucose, and blood pressure) and 4 Psychological Health (Depression, Self-reported mental health, Social Network Index (SNI), Acculturation score). The overall LC12score, Health Factors Score, Health Factors Score, and Psychological Health were calculated as the unweighted average of the 12 metrics. Participants with a LC12score of 80–100 were considered high CVH; 50–79, moderate CVH; and 0–49 points, low CVH.

**Figure 2.**
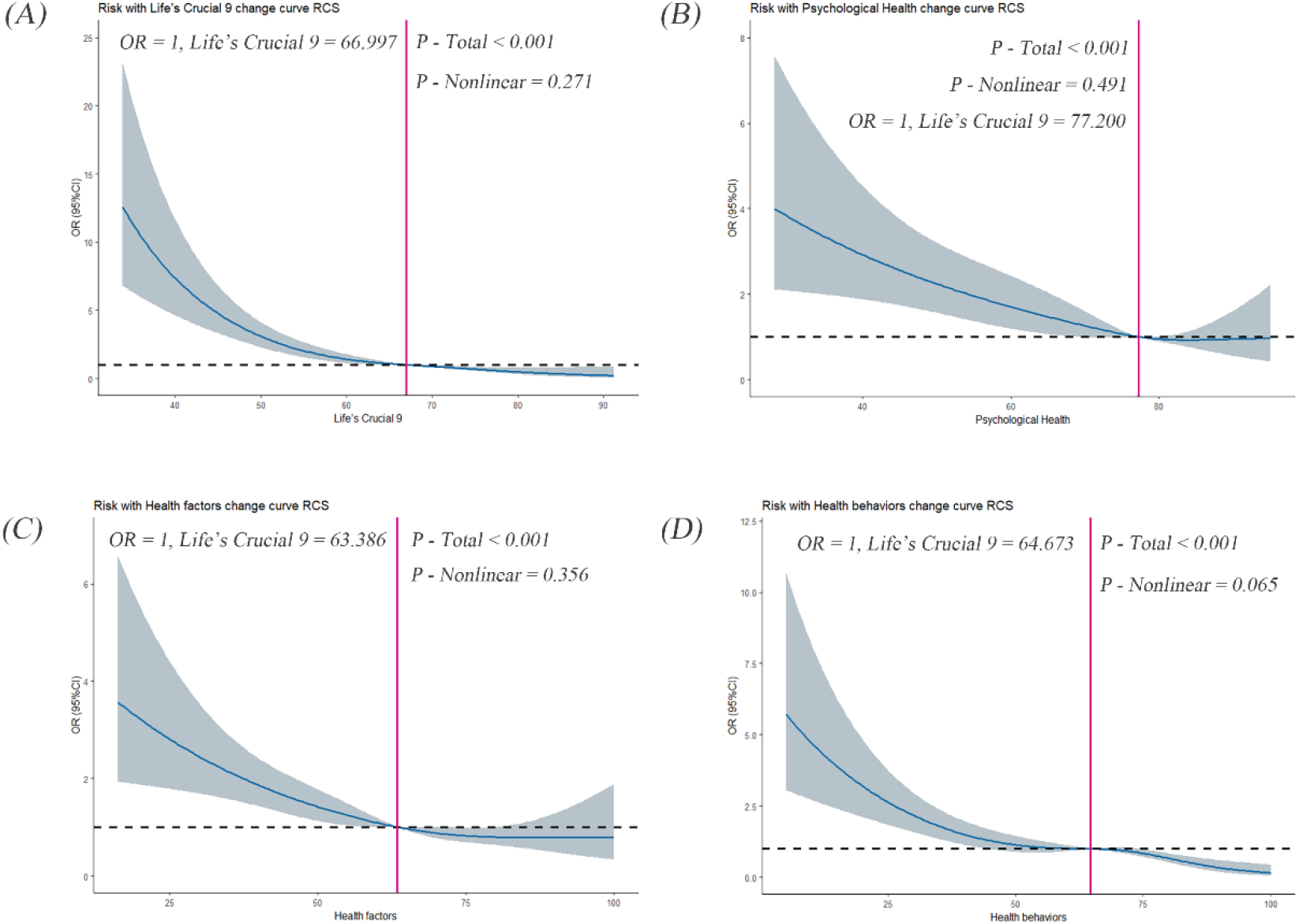
Dose–response relationships between (A), Life’s Crucial 9 (B), Psychological Health (C), Health behaviors (D), Health factors and stroke. OR (95% CI) (shaded areas) were adjusted for Sex, Age, Race, Family income-to-poverty ratio (PIR), Education levels. Vertical red solid lines indicate the minimal threshold for the beneficial association with estimated OR = 1. OR, odds ratio.

### 3.4 Sensitivity analysis

We performed several sensitivity analyses. As shown in Appendix **Table 1**, when the racial grouping (white people, black people) is reset, LC12, Psychological Health, Health behaviors, and Health factors will still reduce Stroke’s risk of disease, which may be protective factors. As shown in Appendix **Table 2**, when the age groups are reset (20– 44, 45–64, 65, and older), LC12, Psychological Health, Health behaviors, and Health factors will still reduce the risk of Stroke and may be protective factors. As shown in Appendix **Table 3**, when Psychological Health and sleep factors are not taken into account, Life’s Simple 7 still reduces Stroke’s risk of disease, which may be a protective factor. As shown in Appendix **Table 4**, Life’s Essential 8 still reduces Stroke’s risk when Psychological Health is not taken into account, possibly as a protective factor.

## 4. Discussion

In this study, we provide two main new findings based on the US general population. First, we constructed the Psychological Health metric and incorporated Psychological Health into the LE8 cardiovascular score model, which was updated with the LC12 cardiovascular health assessment model. In addition, this study adds significant evidence of a relationship between CVH and stroke by using LC 12 as a definition of CVH. We found that LC12 and its components (Psychological Health, Health behaviors, Health factors) are considered to be strong correlates of stroke, and that maintaining a high level of LC12 contributes to reducing the risk of developing stroke.

Numerous studies have explored the correlation between CVH scores (LE8, LS7) and stroke incidence. In both African American and Caucasian American populations, a higher status of LS7 scores has been linked to a decreased likelihood of experiencing a stroke(15). Research has indicated that the lifetime risk of stroke is influenced by genetic predisposition and cardiovascular fitness levels. Higher LS7 levels have been shown to partially mitigate a high genetic risk and lower the overall risk of stroke(42). However, with the replacement of LS7 by LE8, the definition of CVH in LS7 may not comprehensively capture current health practices and behaviors. Studies have identified shortcomings in the quantification of indicators and have suggested sleep health as the eighth component of CVH in LE8(43). LE8 improves the sensitivity of the score to differences between individuals and groups by improving the method of CVH quantification(14). A study from UK Biobank reported that high CVH levels, as defined by LE8, were associated with significantly lower risk of CHD, stroke, and CVD(6), and individuals with lower LE8 scores may have experienced more adverse cardiovascular outcomes (ischemic heart disease, myocardial infarction, stroke, and heart failure)(16). In a cohort study in China, the CVH score assessed by the LE8 index was found to be significantly predictive of future stroke risk and arterial stiffness status and was more significant in young adults and middle-aged adults than in older adults(3). The LE8 has merit as the current mainstream metric for evaluating cardiovascular health. However, the American Heart Association recognizes that Psychological Health plays an important role in achieving optimal and equitable cardiovascular health and has included Psychological Health in its goals for future work(4, 44). Therefore, we developed the LC12 indicators based on the LE8 to assess cardiovascular health. These indicators are more comprehensive measures of Psychological Health, Health behaviors, Health factors, and other factors. Overall, the above studies provide evidence for the association between Health behaviors, Health factors and stroke. The evidence for Psychological Health and Stroke is an important part of the discussion.

Few studies have focused on the relationship between Psychological Health problems and stroke in the general population. Conventional wisdom suggests that negative emotions and Psychological Health are not related to physical health, but there is now growing evidence that they are closely related. For example, one study found that anxiety, stress, and depression were associated with increased cases of stroke(45-47). Furthermore, results of a meta-analysis showed that depression was positively associated with the risk of stroke in adults(45), and that the risk of a first stroke associated with depression was tripled even after adjusting for confounders(48). At the same time, studies have shown that social networks have an important role in neurological disorders, with positive effects on acute stroke care and stroke rehabilitation(49-50), and that smaller social networks are strong predictors of stroke in women at high risk(31). Tomaka et al. found a correlation between social networks and support measures and stroke, and they noted a relationship between stroke and self-reported loneliness and family support(51). On a mechanistic level, poor social connections may lead to increased sympathetic nervous system reactivity to stress, enhanced neurohormonal activation (e.g., elevated cortisol levels), and impaired immune function. This condition may increase the risk of infection and inflammation(52-54). Our findings are similar to previous studies that have used the SNI and other social network metrics to describe the size of an individual’s socialization and the frequency of social contacts. Thus, SNI scores can serve as an important predictor of stroke events(31). Poor self-reported Psychological Health status predisposes to mental illness, the expected risk of psychologically unhealthy days increased over time, and participants with mental illness were more likely to have a stroke(35, 55). In addition, participants with stroke were associated with an elevated risk of self-reported mentally unhealthy days(32). In contrast, acculturation poses a significant risk for stroke(37). Research indicates that immigrant populations experience acute stroke more frequently and more severely in younger individuals(37). Immigrant populations have a higher likelihood of disability following ischemic stroke compared to long-term residents, potentially influenced by factors such as stroke severity, utilization of palliative care, and proficiency in language(56-57). Therefore, there is a need for enhanced focus on rehabilitation and care for immigrant populations. Tailored care and treatment strategies should be developed to mitigate disability risk upon hospital discharge and enhance rehabilitation outcomes for this demographic.

All the aforementioned studies have indicated the significant impact of Psychological Health on stroke risk, with poor Psychological Health status increasing the likelihood of ischemic stroke(22). Elevated mood swings have a strong correlation with stroke and ischemic stroke(58). Hence, during a CVH assessment, the LC12 [4 Psychological Health (Depression, Self-perceived mental health, Social Network Index (SNI), Acculturation score), 4 Health Behaviors (diet, physical activity, nicotine exposure, and sleep duration) and 4 Health Factors (body mass index [BMI], non-high-density-lipoprotein cholesterol, blood glucose, and blood pressure)] could serve as reliable indicators of cardiovascular disease and demonstrate a strong association with stroke risk.

This study also has some strengths. A nationally representative sample of U.S. adults was used, which allows the findings to be generalized to a broader population. We constructed an index LC12 that comprehensively evaluates cardiovascular health, which compensates for the lack of consideration of Psychological Health in the evaluation of CVH. Furthermore, we explored the dose-response relationship between CVH and stroke and identified the minimum threshold for a beneficial association. There are limitations to this study as well. Firstly, it was carried out in a developed nation that might experience healthy volunteer bias, where those willing and able to participate tended to be healthier than the general population. Secondly, health behavior indicators were assessed using self-report questionnaires, which are prone to measurement errors. Moreover, we did not delve into detailed stroke categorization, missing distinctions between various diagnoses with different causes and stroke severities. Lastly, despite controlling for various potential confounders, the cross-sectional design restricted us from making causal or temporal inferences about the relationship between LC12 and stroke risk. Further optimization of the current scoring methodology is necessary. In future research, we aim to gather more comprehensive indicators of Psychological Health (e.g. anxiety, suicidal thoughts, tension) and develop Psychological Health assessment tools using latent category analysis. This approach will offer a more thorough evaluation of CVH in the psychological aspect. Given the constraints of the present study, caution is advised when interpreting these results, and additional research is warranted to validate our findings.

## 5. Conclusion

Overall, higher levels of Health behaviors, Health factors are known to be protective factors for cardiovascular health, and our constructed metrics for evaluating psychological factors (Psychological Health) were strongly associated with CVH evaluation metrics [LC12 (Psychological Health, Health behaviors, Health factors)] are strongly associated with the risk of developing stroke, and individuals will continue to benefit from cardiovascular health when LC12 is maintained at a high level.

## Data Availability

http://ww w.cdc.go/n chs/nhanes.htm

https://www.cdc.gov/nchs/nhanes.htm

## 8. Ethics approval and consent to participate

The surveys were approved by the NCHS Research Ethics Review Board (Protocol #2011-17). All methods were performed in accordance with the relevant guidelines and regulations (Declaration of Helsinki). Informed consent was obtained from all subjects and/or their legal guardian(s).

## 9. Availability of data and materials

All data entered into the analysis were from NHANES, which is publicly accessible to all. All data were openly available, and the ethical clearance certificate and informed consent form were published on the official database website, so there was no need to apply for additional ethical certificates for this study. NHANES has received NCHS Ethics Review Board (ERB) Approval. We show the direct web link to the database in the manuscript: NHANES (http://www.cdc.go/nchs/nhanes.htm).

## 10. Authors’ contributions

Conception and design: Ruoyu Gou, Yufan Gou. Analysis and interpretation of the data: Ruoyu Gou. Drafting of the article: Ruoyu Gou, Yufan Gou, Danni Dou. Critical revision of the article for important intellectual content: Ruoyu Gou, Yufan Gou, Danni Dou, Guanghua Li. Final approval of the article: Ruoyu Gou, Guanghua Li. Statistical expertise: Ruoyu Gou.

## 11. Acknowledgments

Not Applicable.

