## Supplementary material for "Life’s Crucial 12: Updating and Enhancing the Life’s Essential 8 of Cardiovascular Health: A proposal from NHANES"

* Correspondence:

Guanghua Li

### Method

### Life's Crucial 12 Construction Elaborated

We propose to consider updating the indicators for Psychological Health(1) from the original eight indicators of Life's Essential 8 to construct an integrated cardiovascular care model (Life's Crucial 12, LC12) to more fully consider inter-individual differences and intra-individual variation. The aim is for researchers, health systems, and policy makers to create standardized tools to measure and monitor CVH in individuals and populations. The evaluation criteria and detailed calculations for the Life's Essential 8 have been reported in previous studies(2-4), and the subquestionnaires for the LE8 score consist of 4 health behaviors (diet, physical activity, nicotine exposure, and sleep time) and 4 health factors. sleep duration) and 4 health factors (body mass index, non-HDL cholesterol, blood glucose, and blood pressure). Briefly, the 8 CVH metrics range from 0 to 100, and the total LE8 score is calculated as the arithmetic mean of the 8 metrics according to the American Heart Association's definitions, with LE8 scores categorized as low (0 to 49), moderate (50 to 79), and high (80 - 100) CVH8. Dietary indicators were assessed by the Healthy Eating Index (HEI) 2015: participants' dietary intake collected from two 24-hour dietary recalls was combined with food pattern equivalency data from the United States Department of Agriculture (USDA) to construct and calculate HEI-2015 scores.16 Self-assessment questionnaires were used to collect information on the frequency and duration of vigorous or moderate physical activity in the past 30 days, smoking status, sleep duration, history of diabetes, medication history, blood pressure, height, and weight measured on physical examination. Body mass index (BMI) was calculated as weight in kilograms divided by the square of height in meters. Blood pressure was determined as the average of 3 consecutive measurements. After fasting for at least 8 hours, a 5 ml blood sample was collected and sent to the central laboratory for determination of complete blood count and biochemical parameters (e.g., lipids, glucose, CRP, and glycosylated hemoglobin).

We followed previous studies and followed the evaluation model of 4 health behaviors (Diet, Physical Activity, Nicotine Exposure, and Sleep Duration), 4 health factors (Body Mass Index, Non-HDL Cholesterol, Blood Sugar, and Blood Pressure) in the Life's Essential 8 score(2).The subquestionnaire of the LC12 score includes 4 health behaviors (Diet, physical activity, nicotine exposure, and sleep duration), 4 health factors (body mass index, non-HDL cholesterol, blood glucose, and blood pressure), and 4 types of Psychological Health (Depression, Self-reported mental health, Social Network Index (SNI), Acculturation score). Acculturation score). The Psychological Health score is the sum of the Depression, Self-reported mental health, Social Network Index (SNI) and Acculturation score, and the Life's Crucial 12 score is the Healthy Behaviors. The Life's Crucial 12 score is the sum of the Health Behavior, Health Factors, and Mental Health scores. In short, the 12 CVH indicators range from 0 to 100, and the total LE8 score is calculated as the arithmetic mean of the 12 indicators, with Life's Crucial 12 scores categorized as low (0-49), medium (50-79), and high (80-100) CVH. In the following paragraphs, the following is elaborated upon Psychological Health (Depression, Self-reported mental health, Social Network Index (SNI), Acculturation score) is defined and scored in detail in the following paragraphs.

**1. Depression**

In the cross-sectional study, we considered depressive states to be a very important part of mental health problems. Depressive symptoms were assessed using the Participant Health Questionnaire (PHQ-9), a nine-item screening tool designed to assess the severity of depressive symptoms. Each item was scored from 0 ("not at all") to 3 ("almost daily") based on the frequency of depressive symptoms(5). Participants' scores were summarized and individuals were categorized into minimal (0-4), mild (5-9), moderate (10-14), or severe (15-27) depression groups(5) (Depression corresponds to Life's Crucial 12 as minimal (100 score), mild (75 score), moderate (50 score), and severe (25 score)).

**2. Social Network Index (SNI)**

We followed previous research in which the SNI consisted of three components: being married (no = 0, yes = 1), attending religious services (once a month or more = 1, otherwise = 0) and sociability ((four or more close friends/relatives = 1), otherwise = 0)(6). The survey design did not allow respondents to individually identify close friends and relatives. Our SNI ranges from 0 - 3 points, with higher values indicating greater social integration(6). For the purposes of the Social Competence Score, a close friend or relative is someone with whom the respondent feels at ease, can talk about personal matters, and can ask for help when needed (Social Integration scores correspond to Life's Crucial 12 on a scale of 0 (25 points), 1 (50 points), 2 (75 points), and 3 (100 points)).

**3. Acculturation index**

A 5-point acculturation score was constructed in NHANES based on country of birth, length of time in the U.S., and language spoken at home. A score of 3 is based on country of birth and length of time in the U.S., i.e., 3 for U.S.-born, 2 for foreign-born with ≥20 years of U.S. residence, 1 for foreign-born with 10 to 19 years of U.S. residence, and 0 for foreign-born with <10 years of U.S. residence. Language spoken at home was scored as 2 points, i.e., 2 points for English only or primarily, 1 point for both languages equally, and 0 points for other languages only or primarily. These scores were added together to obtain a 5-point acculturation score, with larger values indicating a higher degree of acculturation(7). In other words, the acculturation score is composed of three indicators: country of birth, length of residence in the United States, and language spoken at home. Language spoken at home (between 0-2 points): 0 (Spanish or favoring Spanish), 1 (divided between Spanish and English used with equal frequency), and 2 (English only or favoring English). Social adjustment scores (between 0 and 3): 0 (foreign-born and resided in the U.S. for <10 years), 1 (foreign-born and resided in the U.S. for 10-19 years), 2 (foreign-born and resided in the U.S. for ≥20 years), and 3 (born in the U.S.). We summed the two, and the acculturation scores ranged from 0-5 (the Social Adjustment Index corresponds to Life's Crucial 12 as 0 (score 0), 1 (score 20), 2 (score 40), 3 (score 60), 4 (score 80), and 5 (score 100)).

**4. Self-reported mental health**

In NHANES 2005-2008, mental health status is measured through a self-report questionnaire (7). We used this indicator to assist in determining whether or not mental health was present. Participants were asked, "Now thinking about {your/SP's} mental health, which includes stress, depression, and problems with emotions, for how many days during the past 30 days was {your/his/her} mental health not good?". Mental ill-health status was categorized as "none" (feeling mentally healthy 0-6 days per month, score calculation: (30-reported days)/30*4/5*100) or "yes" (feeling mentally unhealthy 7-30 days per month, score calculation: (30 - number of days reported)/30*1/5*100).

**Appendix Table 1**. Multiple logistic regression models of Life's Crucial 9 with stroke for participants.

| Parameter | model 1 | | Model 2 | | Model 3 | |
|---|---|---|---|---|---|---|
| | **OR (95%CI)** | ***P-value*** | **OR (95%CI)** | ***P-value*** | **OR (95%CI)** | ***P-value*** |
| **Life's Crucial 12** [a] | | | | | | |
| Low | **ref** | | **ref** | | **ref** | |
| Moderate | 0.542(0.289,1.015) | 0.055 | 0.431(0.228, 0.814) | 0.012 | 0.460(0.250, 0.846) | 0.016 |
| High | 0.257(0.076,0.867) | 0.030 | 0.184(0.054, 0.633) | 0.010 | 0.220(0.062, 0.783) | 0.023 |
| *P-trend* | <0.001 | | <0.001 | | <0.001 | |
| **Psychological Health** [a] | | | | | | |
| Low | **ref** | | **ref** | | **ref** | |
| Moderate | 0.621(0.366,1.052) | 0.074 | 0.525(0.286, 0.963) | 0.039 | 0.571(0.315, 1.036) | 0.063 |
| High | 0.388(0.201,0.750) | 0.007 | 0.348(0.178, 0.682) | 0.004 | 0.385(0.188, 0.788) | 0.013 |
| *P-trend* | <0.001 | | <0.001 | | <0.001 | |
| **Health Behaviors** [a] | | | | | | |
| Low | **ref** | | **ref** | | **ref** | |
| Moderate | 0.755(0.547,1.042) | 0.084 | 0.706(0.503, 0.991) | 0.045 | 0.730(0.517, 1.030) | 0.070 |
| High | 0.346(0.185,0.646) | 0.002 | 0.316(0.170, 0.588) | <0.001 | 0.332(0.178, 0.619) | 0.002 |
| *P-trend* | <0.001 | | <0.001 | | <0.001 | |
| **Health Factors** [a] | | | | | | |
| Low | **ref** | | **ref** | | **ref** | |
| Moderate | 0.487(0.351,0.674) | <0.001 | 0.537(0.399, 0.723) | <0.001 | 0.547(0.403, 0.742) | <0.001 |
| High | 0.320(0.180,0.569) | <0.001 | 0.502(0.287, 0.879) | 0.019 | 0.517(0.291, 0.919) | 0.027 |
| *P-trend* | <0.001 | | <0.001 | | <0.001 | |

Model 1: Did not adjust any covariates.

Model 2: Adjusted for age, gender, race.

Model 3: Adjusted for age, gender, race, PIR (family income to poverty ratio), education.

[a] Categorical variable.

Race (white people, black people).

Life's Crucial 12, The components of Life's Crucial 12 include diet, physical activity, nicotine exposure, sleep health, body mass index, blood lipids, blood glucose, blood pressure, Depression, Self-reported mental health, Social Network Index (SNI), and Acculturation score. Each metric has a new scoring algorithm ranging from 0 to 100 points, allowing generation of a new composite cardiovascular health score (the unweighted average of all components) that also varies from 0 to 100 points. LC12scoring algorithm consists of 4 Health Behaviors (diet, physical activity, nicotine exposure, and sleep duration), 4 Health Factors (body mass index [BMI], non-high-density-lipoprotein cholesterol, blood glucose, and blood pressure) and 4 Psychological Health (Depression, Self-reported mental health, Social Network Index (SNI), Acculturation score). The overall LC12score, Health Factors Score, Health Factors Score, and Psychological Health were calculated as the unweighted average of the 12 metrics. Participants with a LC12score of 80–100 were considered high CVH; 50–79, moderate CVH; and 0–49 points, low CVH.

**Appendix Table 2**. Multiple logistic regression models of Life's Crucial 9 with stroke for participants.

| Parameter | model 1 | | Model 2 | | Model 3 | |
|---|---|---|---|---|---|---|
| | **OR (95%CI)** | ***P-value*** | **OR (95%CI)** | ***P-value*** | **OR (95%CI)** | ***P-value*** |
| **Life's Crucial 12** [a] | | | | | | |
| Low | **ref** | | **ref** | | **ref** | |
| Moderate | 0.542(0.289,1.015) | 0.055 | 0.431(0.228, 0.816) | 0.013 | 0.460(0.247, 0.856) | 0.018 |
| High | 0.257(0.076,0.867) | 0.030 | 0.188(0.055, 0.643) | 0.011 | 0.222(0.062, 0.803) | 0.025 |
| *P-trend* | <0.001 | | <0.001 | | <0.001 | |
| **Psychological Health** [a] | | | | | | |
| Low | **ref** | | **ref** | | **ref** | |
| Moderate | 0.621(0.366,1.052) | 0.074 | 0.536(0.290, 0.993) | 0.048 | 0.580(0.315, 1.066) | 0.075 |
| High | 0.388(0.201,0.750) | 0.007 | 0.345(0.175, 0.680) | 0.004 | 0.377(0.182, 0.784) | 0.013 |
| *P-trend* | <0.001 | | <0.001 | | <0.001 | |
| **Health Behaviors** [a] | | | | | | |
| Low | **ref** | | **ref** | | **ref** | |
| Moderate | 0.755(0.547,1.042) | 0.084 | 0.707(0.511, 0.977) | 0.037 | 0.730(0.522, 1.020) | 0.063 |
| High | 0.346(0.185,0.646) | 0.002 | 0.306(0.166, 0.563) | <0.001 | 0.319(0.172, 0.591) | 0.002 |
| *P-trend* | <0.001 | | <0.001 | | <0.001 | |
| **Health Factors** [a] | | | | | | |
| Low | **ref** | | **ref** | | **ref** | |
| Moderate | 0.487(0.351,0.674) | <0.001 | 0.562(0.416, 0.760) | <0.001 | 0.572(0.420, 0.780) | 0.002 |
| High | 0.320(0.180,0.569) | <0.001 | 0.497(0.279, 0.883) | 0.020 | 0.509(0.281, 0.922) | 0.029 |
| *P-trend* | <0.001 | | <0.001 | | <0.001 | |

Model 1: Did not adjust any covariates.

Model 2: Adjusted for age, gender, race.

Model 3: Adjusted for age, gender, race, PIR (family income to poverty ratio), education.

[a] Categorical variable.

Age (20–44, 45–64, 65, and older).

Life's Crucial 12, The components of Life's Crucial 12 include diet, physical activity, nicotine exposure, sleep health, body mass index, blood lipids, blood glucose, blood pressure, Depression, Self-reported mental health, Social Network Index (SNI), and Acculturation score. Each metric has a new scoring algorithm ranging from 0 to 100 points, allowing generation of a new composite cardiovascular health score (the unweighted average of all components) that also varies from 0 to 100 points. LC12scoring algorithm consists of 4 Health Behaviors (diet, physical activity, nicotine exposure, and sleep duration), 4 Health Factors (body mass index [BMI], non-high-density-lipoprotein cholesterol, blood glucose, and blood pressure) and 4 Psychological Health(Depression, Self-reported mental health, Social Network Index (SNI), Acculturation score). The overall LC12score, Health Factors Score, Health Factors Score, and Psychological Health were calculated as the unweighted average of the 12 metrics. Participants with a LC12score of 80–100 were considered high CVH; 50–79, moderate CVH; and 0–49 points, low CVH.

**Appendix Table 3**. Multiple logistic regression models of Life's Simple 7 with stroke for participants.

| **Parameter** | **model 1** | | **Model 2** | | **Model 3** | |
|---|---|---|---|---|---|---|
| | **OR (95%CI)** | ***P-value*** | **OR (95%CI)** | ***P-value*** | **OR (95%CI)** | ***P-value*** |
| **Life's Simple 7** [a] | | | | | | |
| Poor | **ref** | | **ref** | | **ref** | |
| Intermediate | 0.327(0.225,0.475) | <0.001 | 0.401(0.268,0.601) | <0.001 | 0.437(0.295,0.647) | <0.001 |
| Ideal | 0.088(0.027,0.291) | <0.001 | 0.132(0.038,0.459) | 0.003 | 0.159(0.046,0.547) | 0.006 |
| *P-trend* | <0.001 | | <0.001 | | <0.001 | |

Model 1: Did not adjust any covariates.

Model 2: Adjusted for age, gender, race.

Model 3: Adjusted for age, gender, race, PIR (family income to poverty ratio), education.

[a] Categorical variable.

Psychological health and sleep not taken into account.

Life's Simple 7 (LS7) framework encompasses various factors that contribute to overall health, including physical activity, smoking habits, BMI, dietary patterns, blood glucose levels, blood pressure, and total cholesterol levels. the entire LS7 score was therefore categorized as being insufficient (0–7), average (8-10), or ideal (11–14).

**Appendix Table 4**. Multiple logistic regression models of Life's Essential 8 with stroke for participants.

| Parameter | model 1 | | Model 2 | | Model 3 | |
|---|---|---|---|---|---|---|
| | OR (95%CI) | *P-value* | OR (95%CI) | *P-value* | OR (95%CI) | *P-value* |
| **Life's Essential 8** [a] | | | | | | |
| Low | **ref** | | **ref** | | **ref** | |
| Moderate | 0.552(0.334,0.912) | 0.022 | 0.542(0.324,0.904) | 0.021 | 0.569(0.338,0.958) | 0.036 |
| High | 0.299(0.092,0.974) | 0.045 | 0.306(0.091,1.033) | 0.056 | 0.358(0.100,1.286) | 0.107 |
| *P-trend* | <0.001 | | <0.001 | | <0.001 | |
| **Health Behaviors** [a] | | | | | | |
| Low | **ref** | | **ref** | | **ref** | |
| Moderate | 0.733(0.506,1.062) | 0.097 | 0.685(0.482,0.972) | 0.036 | 0.722(0.491,1.061) | 0.092 |
| High | 0.336(0.170,0.666) | 0.003 | 0.288(0.149,0.556) | <0.001 | 0.313(0.158,0.619) | 0.002 |
| *P-trend* | <0.001 | | <0.001 | | <0.001 | |
| **Health Factors** [a] | | | | | | |
| Low | **ref** | | **ref** | | **ref** | |
| Moderate | 0.550(0.389,0.777) | 0.002 | 0.605(0.422,0.868) | 0.009 | 0.618(0.427,0.893) | 0.014 |
| High | 0.381(0.200,0.726) | 0.005 | 0.560(0.289,1.085) | 0.082 | 0.562(0.283,1.118) | 0.095 |
| *P-trend* | <0.001 | | <0.001 | | <0.001 | |

Model 1: Did not adjust any covariates.

Model 2: Adjusted for age, gender, race.

Model 3: Adjusted for age, gender, race, PIR (family income to poverty ratio), education.

[a] Categorical variable.

Psychological health not taken into account.

Life's Essential 8, The components of Life's Essential 8 include diet (updated), physical activity, nicotine exposure (updated), sleep health (new), body mass index, blood lipids (updated), blood glucose (updated), and blood pressure. Each metric has a new scoring algorithm ranging from 0 to 100 points, allowing generation of a new composite cardiovascular health score (the unweighted average of all components) that also varies from 0 to 100 points. LE8 scoring algorithm consists of 4 Health Behaviors (diet, physical activity, nicotine exposure, and sleep duration) and 4 Health Factors (body mass index [BMI], non-high-density-lipoprotein cholesterol, blood glucose, and blood pressure). The overall LE8 score, Health Factors Score and Health Factors Score were calculated as the unweighted average of the 8 metrics. Participants with a LE8 score of 80–100 were considered high CVH; 50–79, moderate CVH; and 0–49 points, low CVH.
